# Development of a multimodal machine-learning fusion model to non-invasively assess ileal Crohn’s disease endoscopic activity

**DOI:** 10.1101/2021.08.29.21262424

**Authors:** Itai Guez, Gili Focht, Mary-Louise C.Greer, Ruth Cytter-Kuint, Li-Tal Pratt, Denise A. Castro, Dan Turner, Anne M. Griffiths, Moti Freiman

## Abstract

**Background and Objective:** Recurrent attentive non-invasive observation of intestinal inflammation is essential for the proper management of Crohn’s disease (CD). The goal of this study was to develop and evaluate a multimodal machine-learning (ML) model to assess ileal CD endoscopic activity by integrating information from Magnetic Resonance Enterography (MRE) and biochemical biomarkers.

**Methods:** We obtained MRE, biochemical and ileocolonoscopy data from the multi-center ImageKids study database. We developed an optimized multimodal fusion ML model to non-invasively assess terminal ileum (TI) endoscopic disease activity in CD from MRE data. We determined the most informative features for model development using a permutation feature importance technique. We assessed model performance in comparison to the clinically recommended linear-regression MRE model in an experimental setup that consisted of stratified 2-fold validation, repeated 50 times, with the ileocolonoscopy-based Simple Endoscopic Score for CD at the TI (TI SES-CD) as a reference. We used the predictions’ mean-squared-error (MSE) and the receiver operation characteristics (ROC) area under curve (AUC) for active disease classification (TI SEC-CD≥3) as performance metrics.

**Results:** 121 subjects out of the 240 subjects in the ImageKids study cohort had all required information (Non-active CD: 62 [51%], active CD: 59 [49%]). Length of disease segment and normalized biochemical biomarkers were the most informative features. The optimized fusion model performed better than the clinically recommended model determined by both a better median test MSE distribution (7.73 vs. 8.8, Wilcoxon test, p<1e-5) and a better aggregated AUC over the folds (0.84 vs. 0.8, DeLong’s test, p<1e-9).

**Conclusions:** Optimized ML models for ileal CD endoscopic activity assessment have the potential to enable accurate and non-invasive attentive observation of intestinal inflammation in CD patients. The presented model will be made available to the community through a dedicated website upon acceptance.

## Introduction

An estimated 1.6 million people in the United States suffer from inflammatory bowel diseases (IBD). Half are believed to have Crohn’s disease (CD) and at least 80,000 are children [1]. While CD can arise within any section of the gastrointestinal tract, it is most prevalent in the small bowel with more than 50% of CD patients having terminal ileum (TI) involvement [2]. CD has a chronic, relapsing, and remitting clinical course. Long-standing inflammation can result in bowel obstruction, stricture, fistula, and/or abscess. In addition, there is an increased risk for small and/or large bowel malignancy in areas of chronic inflammation [3]. Hence, regular observation of intestinal inflammation is essential throughout the life of CD patients.

The Simple Endoscopic Score for CD (SES-CD) [4], assessed by an ileocolonoscopy exam, is considered the most established score for CD endoscopic activity evaluation due to its simplicity and strict scoring convention which results in “excellent” inter-rater and intra-rater agreement [5, 6, 7]. However, ileocolonoscopy is an invasive procedure which requires anesthesia and carries non-negligible risk of perforation [8]. This risk is particularly significant for patients with CD diagnosed at a young age due to required life-long monitoring. Therefore, establishing a non-invasive approach for CD endoscopic activity assessment is urgently needed.

Magnetic resonance enterography (MRE) has emerged during the past two decades as an imaging modality that has the potential to enable non-invasive assessment of CD activity. It has been increasingly used for evaluating CD inflammatory activity, especially for the pediatric population [9, 10]. However, objective MRE-based assessment of CD activity remains an unmet need. Radiologists inter-rater agreement is far from perfect [11, 12], and standardization of assessment parameters for CD is still evolving [13, 14].

Multiple MRE indices were proposed to standardize MRE-based assessment of CD activity [14, 15]. Examples include the Magnetic Resonance Index of Activity (MaRIA) [16], the Pediatric Inflammatory Crohn’s MRE Index (PICMI) [13] and the MRE global score (MEGS) [17, 18], among others. Recently, Turner et al. suggested to non-invasively evaluate CD TI activity from MRE data in pediatric clinical trials with a simple linear-regression (LR) model based on the MaRIA index [19].

Nonetheless, previous studies demonstrated only a moderate correlation between MRE indices and the SES-CD [20, 15]. The moderate correlation may be attributed, at least in part, to the utilization of classical linear models to characterize the correlation between MRE-based variables and endoscopic activity. Such models may be limited in their ability to determine complex and non-linear correlations.

Furthermore, these MRE indices only utilize radiological information, effectively ignoring additional sources of information. Specifically, they disregard biochemical biomarkers such as C-Reactive Protein (CRP) and Fecal Calprotectin (FC) which are known to be indicative of CD severity [21, 22]. However, integration of information from different modalities in a single, comprehensive CD activity index, is challenging when using classical linear models.

In contrast, non-linear machine learning (ML) models demonstrated their potential in leveraging complex non-linear correlations between the input variables and outcomes [23, 24, 25]. Specifically, multimodal ML fusion models combining medical imaging and clinical records improved disease assessment in healthcare tasks [26].

The main goal of this work was, therefore, to develop and evaluate a multimodal ML fusion model combining radiological information and bio-chemical biomarkers for the non-invasive prediction of ileal CD endoscopic activity. To the best of our knowledge, we are the first to propose a multimodal fusion model which combines MRE biomarkers along with biochemical biomarkers, such as CRP and FC, to assess ileal CD endoscopic activity. A secondary goal was to determine the minimal set of MRE-based variables and biochemical biomarkers for ileal CD endoscopic activity assessment.

## Methods

### Ethical approval

The ImageKids study was approved by the Helsinki committee of the Israeli Ministry of Health. All sites obtained research ethics board approval prior to recruitment. Written informed consent was obtained from each participant prior to enrollment in the study. In case of minors, parental or guardian consent was obtained (NCT#01881490 [27]).

Our study used an anonymized version of the Imagekids study database with all identifying elements related to patient privacy removed. Therefore, no additional approval of the ethics committee was required.

### Data collection

This is a substudy of the ImageKids study (NCT#01881490 [27]) in which a total of 240 children, who met the eligibility criteria of being 6 to 18 years old and having an established diagnosis of CD, were enrolled. The data were collected from 22 pediatric IBD centers in North America, Europe, Australia, and Israel [28].

All participants underwent an ileocolonoscopy followed by an MRE within 14 days without any change in treatment. Explicit demographic and clinical data were collected at the time of ileocolonoscopy, including serum biochemical tests and stool samples.

For the purpose of the current study we used only patients with a TI SES-CD which had all features available for the development and validation of our model.

### MRE and radiological assessment

MRE sequences and the acquisition system were standardized across centers [28]. Each MRE protocol included the following sequences: a localizer sequence, a motility sequence in the coronal plane, a series of coronal and/or axial rapid T2-weighted sequences, pre- and post-intravenous gadolinium injection T1-weighted gradient echo sequences, and a diffusion-weighted imaging (DWI) sequence. The intravenous antispasmodic agent (glucagon or hyoscine butylbromide) was administered following the motility sequence. MRE imaging was performed without conducting an enema as it is less feasible in pediatrics.

Centralized reading of each MRE was performed by two independent radiologists, highly experienced in pediatric IBD. The radiologists were blinded to the biochemical and endoscopic data. They recorded imaging biomarkers including items in the MaRIA [16] and the PICMI indices [13] along with the length of the diseased segments, among others. The TI was defined, for the purpose of the Imagekids study, as the first 10 cm segment of the ileum measured from the ileocecal valve. In cases of disagreement between the two radiologists, a consolidated consensus was achieved using a structured voting system as described by Focht et al. [27, 13].

### Endoscopic report and gastroenterologist assessment

Local site gastroenterologists, who were un-blinded to clinical data, performed an ileocolonscopy on each patient and used the Simple Endoscopic score for CD (SES-CD) for assessment [4]. The SES-CD was calculated for each bowel segment separately. A final score was computed as the sum of the segment scores. For the purpose of the current study we used only the TI SES-CD. The SES-CD is not recommended for the assessment of endoscopic activity for patients after bowel surgery [29], therefore patients with previous bowel surgery were excluded from this study.

### TI Normalized biochemical biomarkers

Biochemical biomarkers such as CRP and FC are indicative of CD severity and have a significant correlation with total SES-CD. However, they are non-specific and cannot provide information specific to the TI. We determined the contribution of the TI to the overall SES-CD by normalizing the overall biochemical biomarker score with the relative fraction of the length of the diseased TI out of the entire length of diseased bowel segments as follows:

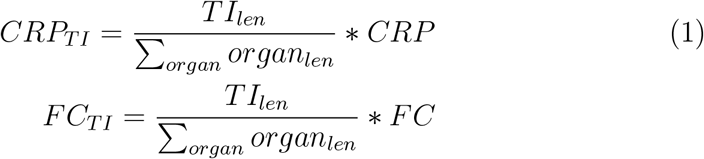

where len is the length of the diseased segment in the specific bowel segment (organ).

### Machine learning model development

Our goal is to approximate the non-linear function that associates the MRE items and biochemical variables to endoscopic activity as assessed by the SES-CD.

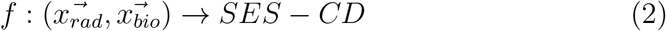

where 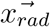 represents the radiological variables and 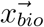 represents the biochemical variables.

We used a Random Forest (RF) model [30] to approximate the function f. The RF model is trained by dividing its development set into various subsets of biomarkers to construct several decision trees. Then, the model aggregates the decisions from the different trees into a final decision [30]. The RF model is suitable for our task as it does not require large amounts of data for training and especially suits an ordinal target such as the TI SES-CD.

### Models’ development and evaluation

We used a stratified 2-fold validation, repeated 50 times with a total of 100 folds, in order to train each algorithm version with a random set of patients used for the development and validation sets. On each repetition, we divided the study cohort into two balanced disjoint sets, each made up from ∼50% of the patients, for the development and validation sets, respectively.

We quantified the contribution of each individual feature to the overall TI SES-CD prediction using the permutation feature importance algorithm [31]. Specifically, we evaluated the decrease in accuracy of an RF model trained with all available features (table 1) on the validation set when randomly shuffling the feature under evaluation across patients data. A larger decrease in model performance indicated a higher importance score.

**Table 1:** Feature Description

| Biomarker | Range | Description |
| --- | --- | --- |
| Wall-thickness | Positive Number (mm) | Measure the thickest part within the segment across all sequences |
| Ulcers | No - 0, Yes - 1 | Mucosal indentation smaller than the full thickness of the bowel wall |
| Mesenteric edema (T2 hyperintensity) | No - 0, Yes - 1 | T2 hyperintense signal of the mesentery adjacent to the same bowel loop assessed |
| Mural edema | No - 0, Yes - 1 | T2 hyperintensity relative to psoas muscle |
| Relative Contrast Enhancement (RCE) | Unbounded number (%) | Increased signal intensity of the bowel wall (WSI) on post-contrast T1-weighted sequences, judged subjectively relative to adjacent normal bowel loops and compared with pre-contrast T1-weighted images. WSI is calculated by the average of three wall enhancement measurements. $RCE = [(WSI_{\text{postgadolinium}} - WSI_{\text{pregadolinium}}) / (WSI_{\text{pregadolinium}})] * 100 * (SD_{\text{noise pregadolinium}} / SD_{\text{noise postgadolinium}})$, where $SD_{\text{noise pregadolinium}}$ corresponds to the average of three SD of the signal intensity measured outside of the body before gadolinium injection, and $SD_{\text{noise postgadolinium}}$ corresponds to the SD of the same noise after gadolinium administration. |
| Wall restricted diffusion | None/Mild - 0, Moderate/Marked - 1 | A subjective measure of hyperintensity on highest b-value trace image compared to normal bowel |
| Comb sign | No - 0, Yes - 1 | Vascular engorgement related to the bowel loop assessed |
| Length of disease | Positive Number (cm) | Radiologist's linear measurement of length of bowel affected |
| C-Reactive Protein (CRP) | Positive Number (mg/L) | Protein made by the liver. Levels in the blood increase when there is a condition causing inflammation somewhere in the body. A CRP test measures the amount of CRP in the blood to detect inflammation due to acute conditions or to monitor the severity of disease in chronic conditions. |
| Fecal Calprotectin (FC) | Positive Number ($\mu\text{g/g}$) | Calprotectin is a protein found in human blood, saliva, cerebrospinal fluid, and urine when some part of the body is inflamed, although it is not always possible to tell the location of the inflammation during testing of these fluids. When detected in the stool, calprotectin has a direct relationship (consequence of neutrophil degranulation) to bowel mucosal damage, characteristic of inflammatory bowel disease. |

We calculated distribution of the feature importance scores over the folds. Then, we selected features with the highest mean importance score for optimized model development.

We assessed the prediction accuracy of the optimized model in comparison to models developed with different input combinations and to a clinically recommended LR model based on the MaRIA index [19, 20] with the ilecolonoscopy-based TI SES-CD as a reference. Specifically, we developed 3 models for comparison with the optimized model as follows.

1. **LR-MaRIA**: the currently recommended clinical standard LR model which is based solely on the original MaRIA index (LR-MaRIA). We inferred the intercept and slope which scales the original MaRIA index according to the given development data [19, 20].
2. **RF-Biochemical**: an RF model based only on non-normalized FC and CRP values as input features.
3. **RF-Biochemical-All**: an RF model with all possible MRE features and normalized clinical biomarkers as input features.

### Statistical analysis

We determined the models’ TI SES-CD prediction accuracy by means of the mean-squared-error (MSE) from the reference endoscopic TI SES-CD over the different folds. Statistical analysis was performed with the Wilcoxon non-parametric test [32] with Bonferroni correction to control the family wise error rate (FWER) [33] in order to determine whether the median validation MSE differed between two given models.

We further evaluated the capacity of the predicted TI SES-CD to distinguish between patients with and without endoscopic CD activity. we used ileocolonoscopy-based endoscopic activity assessment by means of the TI SES − CD < 3 as a reference [20]. We modified the models outputs to predict a score in the range of 0-1 by dividing the prediction by the maximum possible SES-CD value of 12.

We used the area under the receiver operating characteristic (ROC) curves aggregated over the folds as the evaluation metric. We determined whether the differences in the area under the ROC curve (AUC) of the different models are statistically significant with the Delong’s test [34].

### Software and hardware

The models were written in Python [version 3.6.4] using the open-source scipy [version 1.5.4] library and the open-source statsmodels [version 0.11.0] library. We used scikit-learn python library [35] RF model implementation to calculate the feature importance scores [31]. All models ran on an Intel i3 CPU.

## Results

### Data selection

Fig. 1 summarizes the patient selection process for the study. A total of 121 patients out of the 240 children in the Imagekids cohort were included in this sub-study, while 119 were excluded due to missing data (5 due to incomplete colonoscopies, 13 due to prior bowel surgeries, 33 due to non-intubated TI and 68 due to missing one or more features [wall restricted diffusion (n=22), RCE (n=22), CRP (n=22), FC (n=35)]).

**Figure 1:**
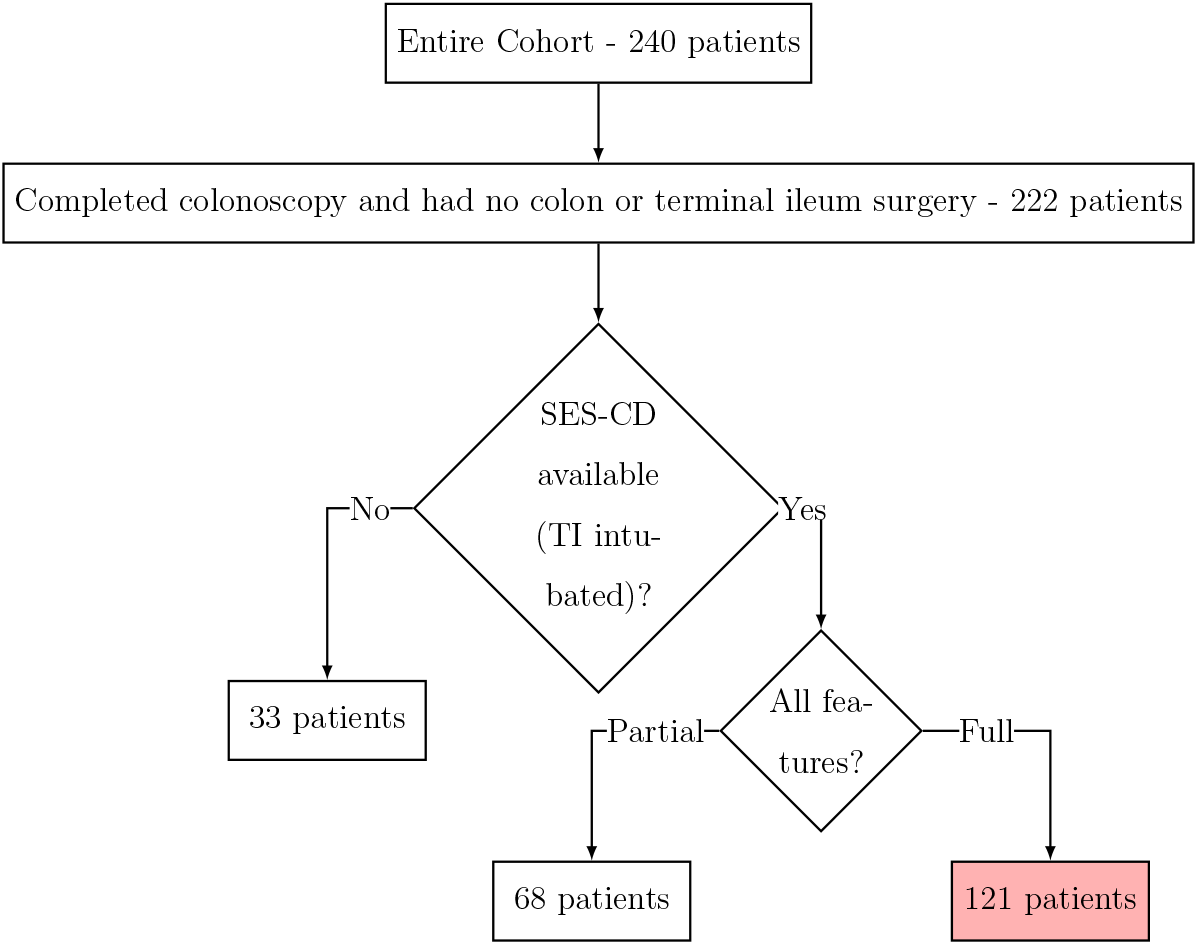
Patients selection from the entire cohort

The detailed SES-CD distribution for the patients included in our study was as follows: 62 subjects (51%) with non-active disease (TI SES-CD<3), 32 subjects (27%) with moderately active disease (3≤TI SES-CD≥ 6) and 27 subjects (22%) with severe disease (TI SES-CD>6). Fig. 2 summarizes the MaRIA scores and the SES-CD for the study’s patient cohort.

**Figure 2:**
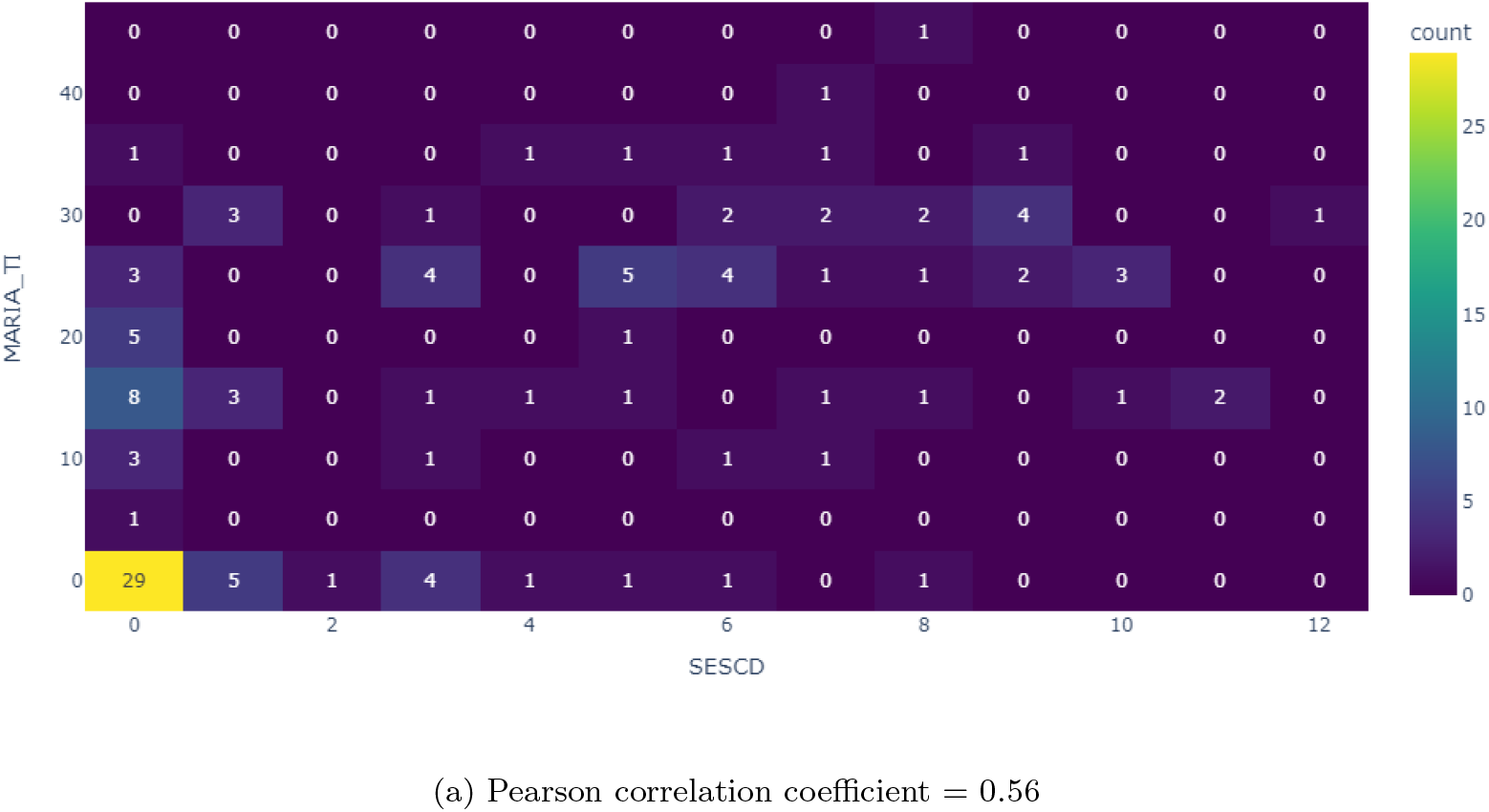
2D histogram of the entire cohort’s endoscopic TI scores and MRE-based MaRIA TI scores

### Feature importance

Fig. 3 summarizes the distribution of the feature importance scores based on the validation accuracy of an RF model trained over the folds with all features as input. The normalized CRP and FC were the two most important features. The MRE features that received the highest scores were the wall thickness and diseased segment length.

**Figure 3:**
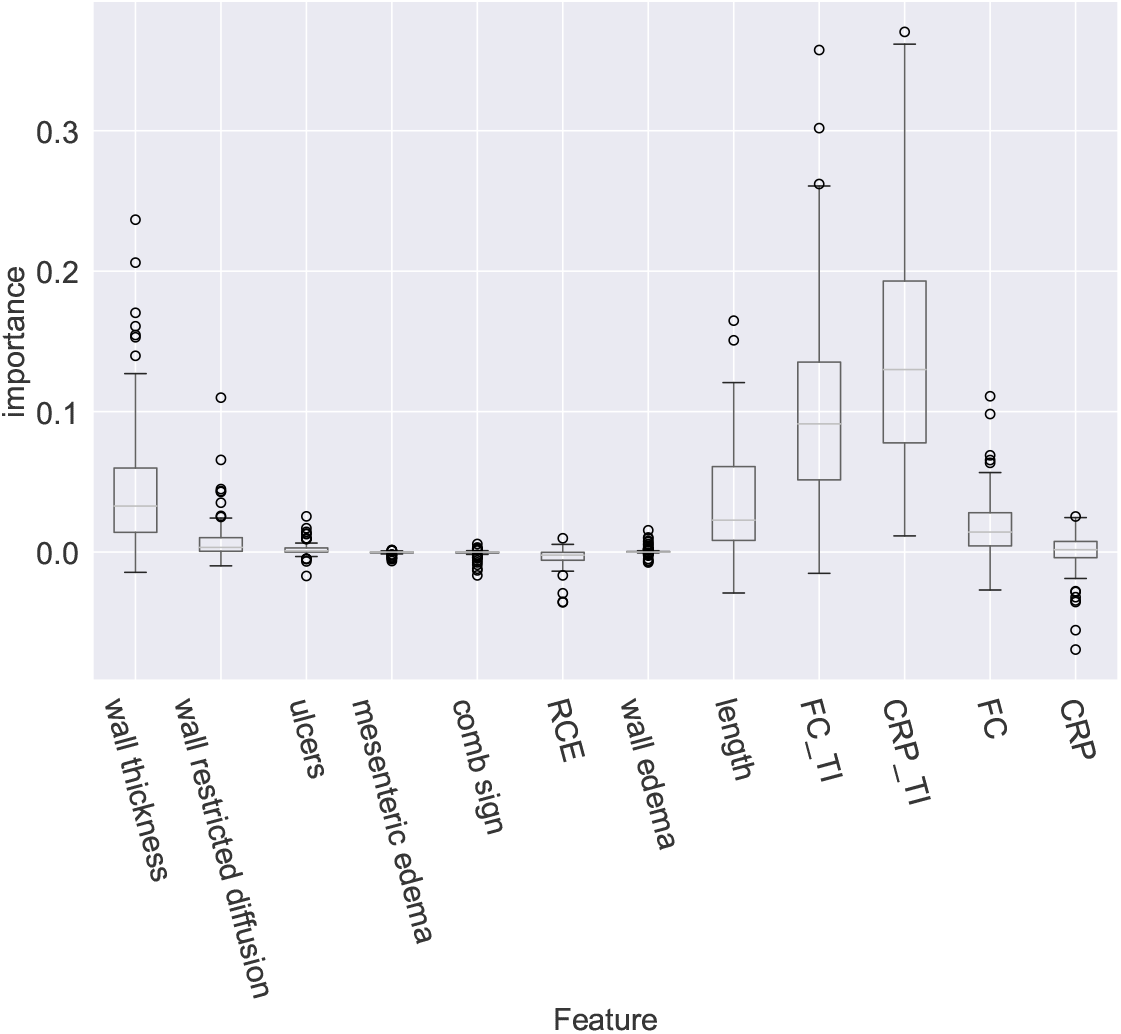
Feature importance distribution over the folds based on feature permutation on the validation-set.

We defined, based on the feature importance results, several feature-sets (table 2) for optimized multimodal fusion models development.

**Table 2:** Feature sets

| **MRE-Set** | **Features** |
| --- | --- |
| Numerical | wall thickness, length of diseased segment |
| MaRIA | wall thickness, RCE, mural edema, ulcers |
| Length | length of diseased segment |
| All | wall thickness, RCE, mural edema,<br>ulcers, wall restricted diffusion, length of diseased segment,<br>mesenteric T2 hyperintensity, comb sign |
| **Biochemical-Set** | **Features** |
| Non-normalized | $CRP, FC$ |
| Normalized | $CRP_{TI}, FC_{TI}$ |

We developed an additional 2 optimized multimodal fusion models as follows:

1. RF-Biochemical-Numerical: consisted of the numerical MRE features enriched with normalized biochemical biomarkers.
2. RF-Biochemical-Length: consisted of only the TI length MRE feature and normalized biochemical biomarkers.

Table 3 summarizes the feature-sets compositions used by all models under evaluation (3 baseline and 2 optimized multimodal fusion models). Table 4 summarizes the RF configuration which was used in developing all the models.

**Table 3:** Feature sets composition

| **Model** | **MRE** | **Biochemical** |
| --- | --- | --- |
| LR-MaRIA | MaRIA | - |
| RF-Biochemical | - | Non-normalized |
| RF-Biochemical-Numerical | Numerical | Normalized |
| RF-Biochemical-Length | Length | Normalized |
| RF-Biochemical-All | All | Normalized |

**Table 4:**
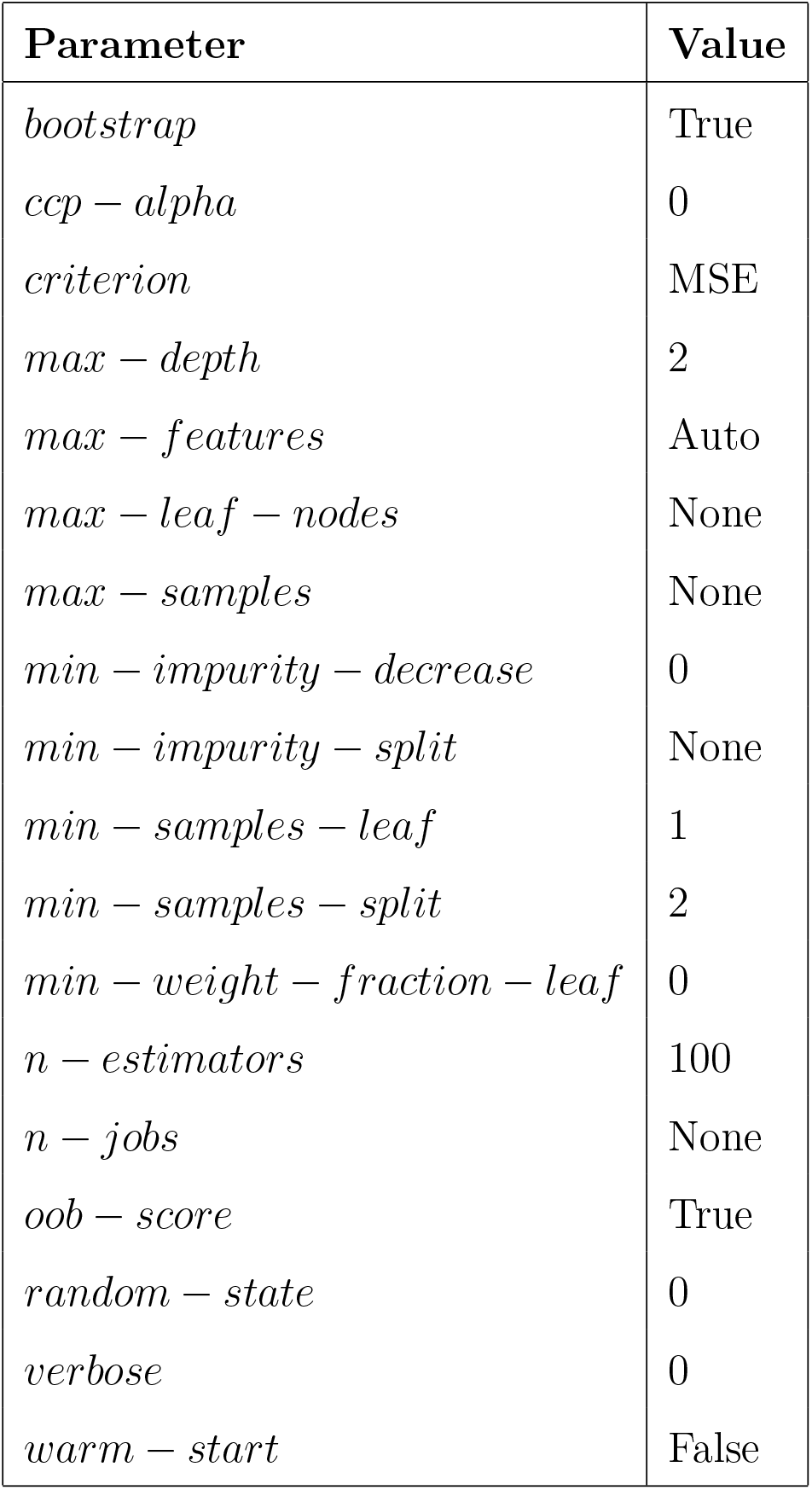
Random Forest configuration

### SES-CD prediction from MRE and biochemical biomarkers on TI intubated patients

Fig. 4 summarizes the distribution of the validation MSE over the folds for each of the models and the calculated p-values.

**Figure 4:**
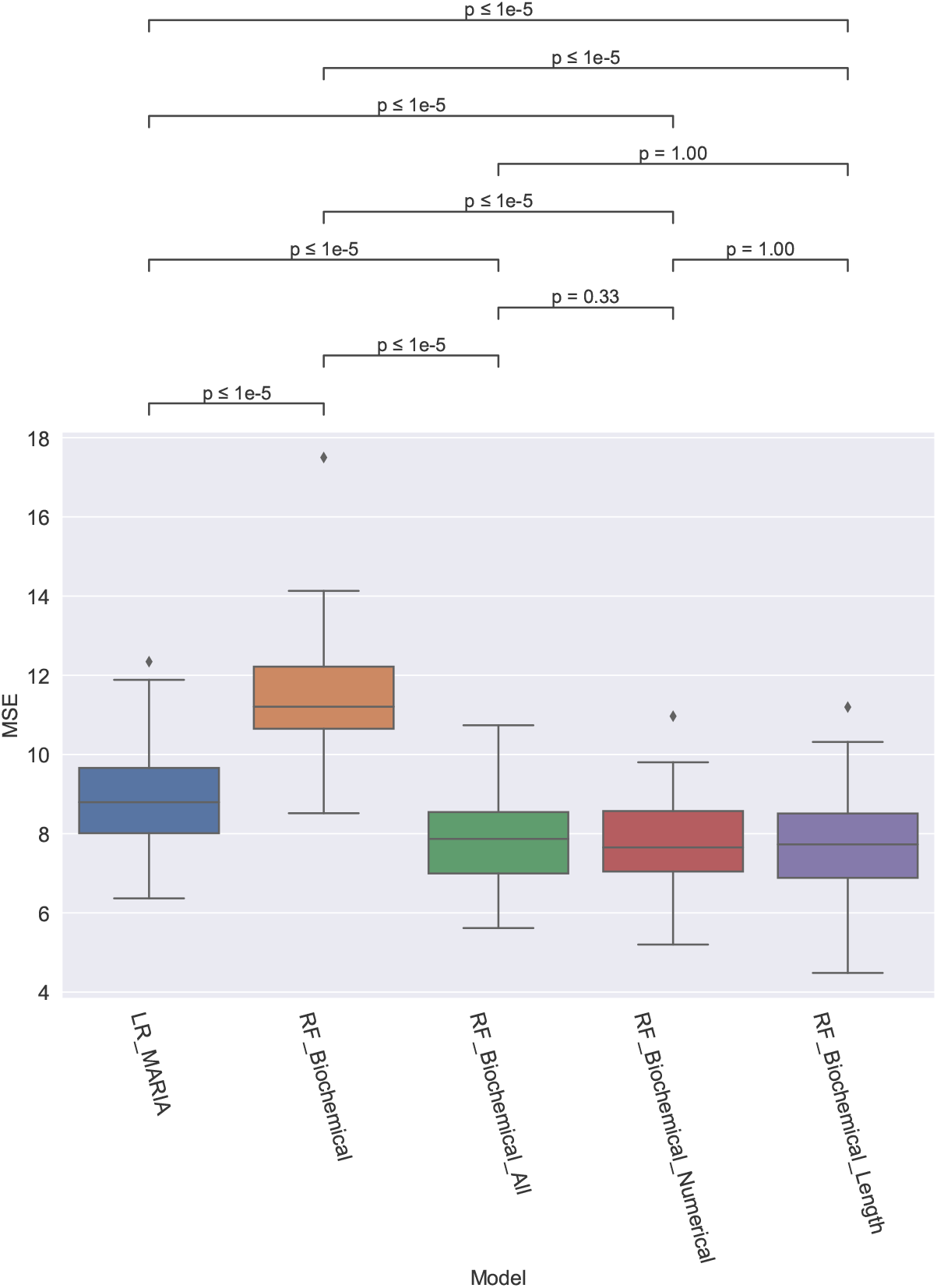
Validation MSE distribution over the folds

All three multimodal fusion models, which were based both on MRE TI features and normalized biochemical features, achieved more accurate results than the baseline models, which were based solely on either MRE features or biochemical features (RF-Biochemical-Length, RF-Biochemical-Numerical, RF-Biochemical-All vs. RF-Biochemical, LR-MARIA, p < 1e − 5). There was no statistically significant difference between the various multimodal fusion models. The baseline model which was based solely on biochemical variables was less accurate than the baseline MRE model (RF-Biochemical vs LR-MARIA, p < 1e − 5).

Fig. 5 presents the averaged ROC curves of the different models for differentiation between patients with non-active disease and patients with active disease according to the reference ileocolonscopy-based SES-CD (SES − CD < 3 [20]).

**Figure 5:**
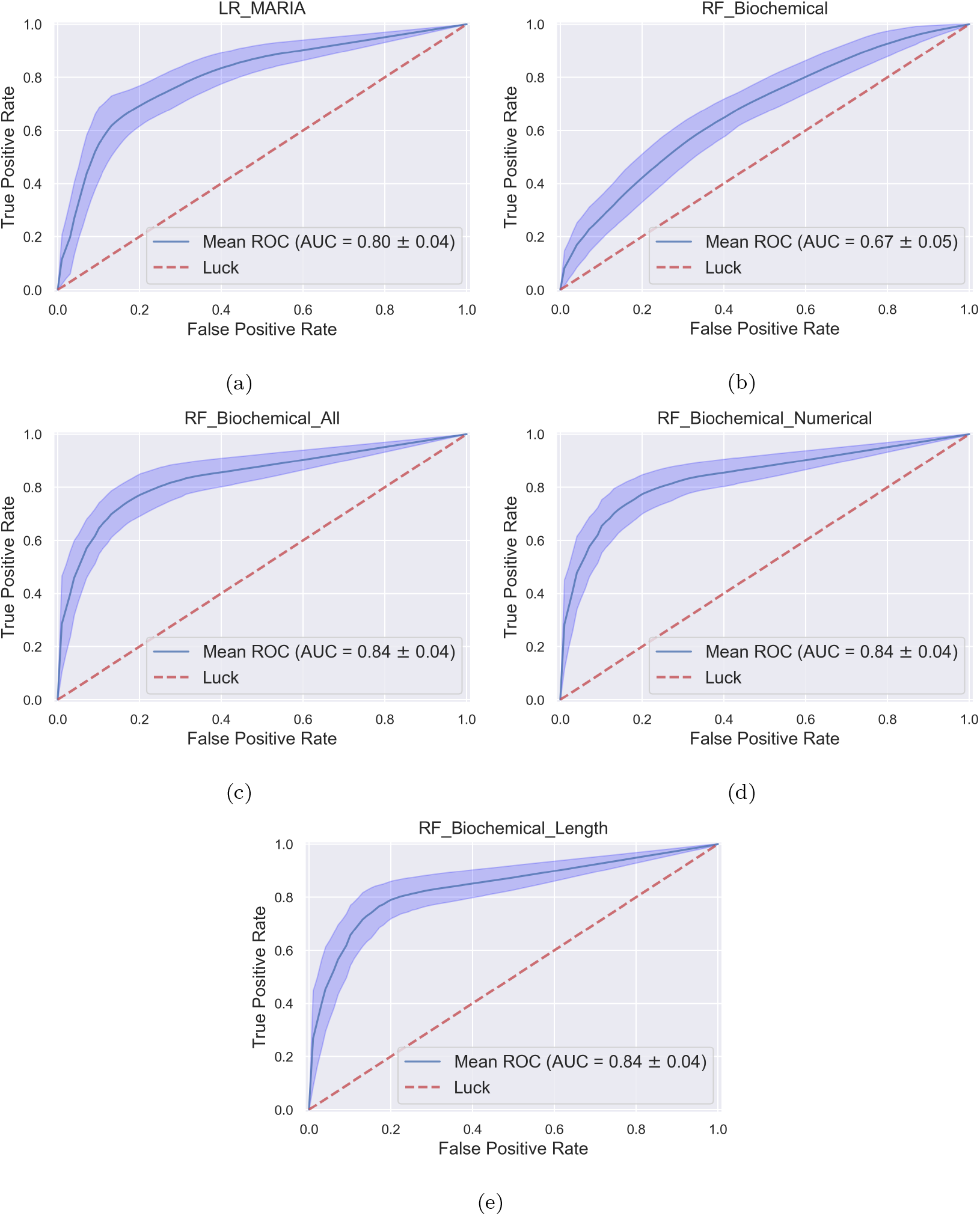
Average ROC curves over the different folds for all models.

All multimodal fusion models (RF-Biochemical-Length, RF-Biochemical-Numerical, RF-Biochemical-All) achieved the highest AUC of 0.84. The difference in the AUC between them and the previously proposed LR-MaRIA was statistically significant (0.84 vs. 0.8, DeLong’s test, p<1e-9).

## Discussion

Multiple non-invasive indices were proposed to objectively evaluate CD activity for clinical trials and routine patient management [36, 37]. However, previously proposed indices relied solely upon either imaging data or biochemical data. Previous studies demonstrated a significant positive correlation between biochemical biomarkers, such as FC and CRP, and active inflammation in both MRE and endoscopy [38, 18]. The biochemical biomarkers reflect molecular data of proteins obtained by immunochemical techniques whereas MRE scans present macrobiology findings. Furthermore, biochemical biomarkers are non-specific and do not indicate disease location. Therefore their role in providing site-specific assessment of CD activity is limited.

Previously, Stawczyk et al. hypothesized that constructing an index consisting of several parameters representing different sources of information (such as MRE and biochemical biomarkers), can lead to a better and more objective CD activity assessment compared to indices based on MRE or biochemical markers alone [39]. However, the development of multimodal fusion models for CD activity assessment that integrate information from multiple sources/modalities is impractical when using classical linear models.

In this study, we demonstrated that a combination of biochemical biomarkers and MRE scans using a non-linear machine-learning model can more accurately reflect the disease phenotype compared to methods that rely on a single source of information.

Although our RF-Biochemical-Length model features are a subset of RF-Biochemical-All and RF-Biochemical-Numerical features, all the models’ performances were broadly similar. Therefore diseased segment length in conjunction with biochemical biomarkers normalized by the relative diseased segment length might be the most important features to evaluate during non-invasive assessments of CD endoscopic activity. Our findings are in agreement with previous studies which demonstrated that the length of the affected segment might be an indicator of the overall burden of CD [18, 14, 40, 17].

The diseased segment length is related both to the transmural inflammation and to the mucosal inflammation while other MRE biomarkers assess mostly transmural inflammation far from the mucosa and therefore cannot be evaluated by an endscopic exam. Yet, the role of the additional MRE biomarkers used in indices such as the MaRIA score remains crucial for transmural active inflammation evaluation.

Further, we demonstrated that non-linear machine learning methods may enable the development of multimodal fusion models for CD activity assessment. Such models intuitively follow the way Gastroenterologists evaluate the disease and assess specific bowel segments.

In addition, our feature selection process reveals the role of the diseased segment length and normalized biochemical biomarkers as the most informative features for non-invasive assessment of ileal CD endoscopic activity. This finding warrants a more comprehensive clinical trial to assess the specific role of length in assessing CD endoscopic activity.

Further, the feature selection process may improve the explainability of our ML-based models to the clinical experts by highlighting the features that were used in models’ predictions. Lastly, a potential benefit of using our proposed model is a reduction of both the number of MR sequences needed, and the number of radiological items needed to be evaluated by the radiologists for the non-invasive prediction of ileal CD endoscopic activity.

In this study, we used the Imagekids study dataset. This dataset is highly heterogeneous and representative of clinical settings (22 sites worldwide, 3 MRI manufacturers); it is rich in patient data from diverse sources. In addition, the Imagekids study dataset includes both MRE and endoscopy examinations conducted within a short time frame, a noteworthy distinction as such closely linked assessments are scarce, especially for the pediatric population. The TI endoscopic activity, used as the model prediction target, is currently considered the primary treatment goal for CD patients in clinical trials. Thus, the models we developed have the potential to provide clinically relevant information to guide clinical trials and patient management. However, additional external validation of our models on larger datasets is warranted before clinical utilization. Nevertheless, such external validation is beyond the scope of the current work.

In the current study we focused on developing ML models for CD endoscopic activity assessment in the TI. Non-invasive assessment of CD endoscopic activity is especially important in the TI as this intestinal segment has the highest prevalence of CD. Further, higher rates of endoscopic TI non-intubation, especially in the pediatric population, necessitate the development of a non-invasive approach to assess ileal CD endoscopic activity. Finally, the development of ML models requires large amounts of data which, in the ImageKids dataset, was sufficiently available for the TI.

While Lee et al. [41] reported a higher AUC against reference TI endoscopic activity than our study for CD assessment from MRE data, several study design differences may have contributed to this difference. First, the Lee et al. study’s cohort was an adult population rather than a pediatric population as in our study. The pediatric population is more challenging to assess as patient movement and imperfect patient bowel preparation occurs more frequently in children and may result in lower quality MRE data. Second, our study used the heterogeneous data of 22 centers worldwide which makes central reading assessment harder and less reliable compared to a single center study which is more homogeneous by nature.

We used the RF model for ML model development. The RF model is most suitable for our task as it can be developed with limited amounts of data, can utilize different types of data as inputs, and can produce ordinal outputs. We also utilized a stratified 2 folds cross-validation approach repeated 50 times to estimate our models reproducibility, and to overcome potential overfitting due to the small dataset that was available for our study.

There are several limitations to our study. First, although the ImageKids study aimed to standardize gastroenterologist evaluations, radiologist readings, MRI machine protocols and the quality of the MREs across sites, some degree of heterogeneity was unavoidable. While this heterogeneity may influence ML model performance, the ImageKids study multicenter enrollment stands as an advantage in reflecting real life variability. Yet, model predictions using biomarkers obtained by radiologists’ readings of MRE data that were not standardized according to the ImageKids MRE protocol should be interpreted with caution.

Second, while the use of central reading is considered an advantage in clinical trials, there is always a risk of introducing reading bias. This was overcome by the double central readings and introducing a third reader in the event of disagreement.

Third, the amount of data that was available for the study was limited. ML models generally require more data for the development stage, in comparison to the data needs of linear models, due to their complexity. Therefore, the presented models’ performances might underestimate the added-value of the ML models for CD endoscopic activity assessment.

Fourth, the length-based approach we used to normalize the biochemical markers assumed that the contribution of each inflamed bowel segment to the biochemical biomarker is proportional to its disease length. However, this assumption is neither validated nor widely accepted. A further investigation of the relative contribution of each inflamed segment to the overall biochemical biomarker is warranted.

Finally, CD can affect the entire GI tract. However, our model development focused only on the TI. Development of models for other bowel segments requires additional data which was not available for this study.

## Conclusion

In this work we addressed the need for a non-invasive assessment of ileal CD by developing a multimodal fusion ML model combining MRE data and biochemical biomarkers. Our optimized model performed the best compared to both the current clinical recommendation of linear models based on the MaRIA score and to ML models developed solely based on radiological variables or biochemical markers and without applying a feature-selection process. Furthermore, the proposed approach can serve in generating clinical hypotheses on the specific roles of the different items used as model inputs in non-invasive CD endoscopic activity assessment.

Our model will be made available to the community through a dedicated website upon acceptance.

## Data Availability

The data that support the findings of this study are available on request from the corresponding author. The data are not publicly available due to restrictions e.g. they contain information that could compromise the privacy of research participants

## Acknowledgments

The ImageKids study was supported by an educational grant from AbbVie who were not involved in any part of the study design, conduct, analysis, or writing. The authors declare no competing non-financial interests but the following competing financial interests: I.G - None. G.F – received last 3 years consultation fee from Abbvie and Lilly M.L.C.G – received in the past 3 years AbbVie investigator-initiated research grant and honoraria, Samsung honoraria. R.C.K - None. L.P - None. D.A.C - None. D.T - received last 3 years consultation fee, research grant, royalties, or honorarium from Janssen, Pfizer, Hospital for Sick Children, Ferring, Abbvie, Takeda, Atlantic Health, Shire, Celgene, Lilly, Roche, ThermoFisher, BMS. A.M.G – received during the past 3 years consultant fees from Abbvie, Amgen, BristolMyersSquibb, Lilly, Janssen, Merck, Pfizer; speaker fees from Abbvie, Janssen, Nestlé; investigator-initiated research grant from Abbvie. M.F - None.

## Author contributions statement

I.G, M.F.: Conception and design, analysis and interpretation of data, drafting the article, and final approval of the version to be published. G.F, M.L.C.G, D.T: Data collection and interpretation of results, revising the paper critically for important intellectual content, and final approval of the version to be published. R.C.K, L.P, D.A.C, A.M.G: Data collection and final approval of the version to be published

